# Maternal health Aggregated Trends can be Misleading: The power of N-of-1 Level Wearable Data Analysis for Personalized Pregnancy Monitoring

**DOI:** 10.1101/2025.10.20.25338363

**Authors:** Tina Behrouzi, Jennifer Yu, Robin Yang, Adrien Boch, Anna Goldenberg, Sarah M. Goodday, Stephen H. Friend

## Abstract

**Background:** Personal digital health technologies (DHTs) enable real-time monitoring of physiological metrics and behavioral data, including HRV, supporting early detection of pregnancy-related conditions and personalized care throughout the perinatal period. While recent studies demonstrate the utility of personal DHTs in tracking pregnancy-related symptoms, they often rely on aggregate statistical methods that overlook individual variability.

**Objective:** To compare aggregate and individual-level analyses of digital health technology (DHT) data for early detection of pregnancy-related conditions, using the comprehensive BUMP dataset to highlight the importance of individual variability and data heterogeneity.

**Methods:** This BUMP study (Jan 2021 – May 2022) analyzed physiological and behavioral metrics, such as heart rate variability (HRV), sleep, and fatigue, in 256 individuals using Oura rings and self-reported surveys. Individual-level (N-of-1) trajectories were evaluated and compared with aggregate results to uncover personal and collective trends. A statistical method was developed to assess the influence of adverse events and severe symptoms, while case studies explored confounding and modifying factors underlying heterogeneity. Comprehensive statistical analysis included the coefficient of determination, Kolmogorov-Smirnov tests, likelihood ratio tests, and Welch’s t-tests, with inter-individual variability flagged based on high-variability thresholds.

**Results:** Results revealed significant variability in HRV, sleep, and fatigue throughout pregnancy. For instance, only 4.76% of individuals had HRV inflection points at the aggregate week 33 inflection, with a 14.24% coefficient of variation. Our analysis found no significant p-values for demographic or pregnancy complication-based subgrouping, suggesting these factors alone do not drive the observed variability. Case studies further highlighted both intra- and inter-individual differences, emphasizing the importance of considering external factors like adverse events and severe symptoms.

**Conclusions:** Our findings show that aggregate wearable data often fails to generalize across populations, oversimplifying pregnancy-related physiological and subjective changes. This simplification can obscure individual trajectories, leading to generalized insights that may not reflect many pregnant women’s experiences. Our results highlight the impact of heterogeneity on pregnancy outcomes, emphasizing the need to move beyond one-size-fits-all models and leverage DHT for personalized care.

## Introduction

Pregnancy is a critical period marked by significant physiological and psychological changes, influencing maternal and fetal health (1). Conditions like gestational hypertension, preeclampsia, preterm birth, and postpartum depression impose significant global health burdens (2). Despite their prevalence, there is still a limited understanding of the underlying factors driving pregnancy outcomes and an essential need for improved support systems to address women’s health during this critical transition (3).

Wearable devices, digital health apps, and other remote smart devices (Personal Digital Health Technologies [DHTs] (4)) are increasingly recognized for their potential to transform healthcare (5,6). DHTs enable the semi-continuous, real-time monitoring of physiological measures and behaviour information, including cardiovascular metrics, activity levels, sleep patterns, and high frequency subjective symptoms (7–9), particularly relevant to pregnancy. These approaches offer an advantage over traditional aperiodic, symptom monitoring methods in capturing subtle individual differences in a remote setting, filling in the gaps between clinic visits, and providing tools that could track key symptoms in populations who lack access to prenatal care (10,11). Harnessing data from diverse DHTs could enhance early detection of pregnancy-related conditions (4,12,13) and enable personalized care throughout the perinatal period (14).

While recent studies have demonstrated the value of personal DHTs in tracking pregnancy-related symptoms and outcomes, they often rely on aggregate statistical methods (14–17) that can overlook individual variability, as a one-size-fits-all approach may not apply effectively to every individual (18). Additionally, some pregnancy studies have faced limitations due to small sample sizes, further reducing the generalizability of their findings. The variability in behaviors and physiological responses further challenges broader applicability of aggregate models. Personal DHTs in pregnancy offer individualized risk monitoring, but recent studies rely on aggregated data instead of N-of-1 analyses. Closing this gap is key to making N-of-1 a standard and maximizing wearable data in future research.

This paper aims to compare the generalizability of personal DHT study results derived from aggregate data with those based on N-of-1 analyses in pregnant individuals using data from a US-based digital health pregnancy study (BUMP study (19)). Three recently published pregnancy digital health studies (15–17) were selected that focused on three key measures; heart rate variability (HRV) (15), sleep (17), and fatigue (16). Aggregated results from these studies were replicated using the BUMP study dataset and were extended by exploring N-of-1 analyses of these key measures. By applying N-of-1 methods, including spline fitting, pregnancy condition analysis, and analyzing certain women in case studies, we aimed to dissect individual differences often overlooked in aggregated approaches.

## Methods

### Study Design

The BUMP study was a participant-centric digital health study that tracked maternal symptoms using wearable devices and smartphone apps from pre-conception, through pregnancy, and up to three months postpartum (19). This analysis focused on three key signals, HRV, sleep (deep sleep and awake time), and fatigue, drawn from multimodal data collected through devices like the Oura ring, and Apple smartwatches. These signals were chosen to capture both objective and subjective measures of health and align with recently published maternal health wearable studies. Nighttime HRV was calculated using the rMSSD method from 5-minute interval data collected by the Oura ring. Awake time and deep sleep were derived from the Oura ring’s 3D accelerometer measurements and 5-minute sleep rankings, respectively. Fatigue was a self-reported feature tracked daily via the BUMP study smartphone app.

### Participants

Out of 431 participants, 275 were included after excluding those with more than 60% missing data, missing delivery information, or fewer than 30 data points. The number of participants in each complication group was as follows: 37 with preterm birth, 42 with gestational diabetes mellitus (GDM), 38 with preeclampsia, 66 with gestational hypertension, 39 with postpartum hemorrhage, 189 with major depression, and 17 were categorized as healthy individuals (as defined as normal weight (BMI <25), age <35 years of old, and no complications during pregnancy and postpartum). Further demographic information is provided in Table S4.

### Data Preprocessing

Details on data collection and study measures are in the Supplementary Methodology. We used aggregated daily summaries from 5-minute interval sleep data, calculating total duration for each sleep stage (deep sleep, awake, REM) across the sleep period. Moreover, HRV was averaged over the entire sleep period. Duplicated time points for objective features were removed. Outliers were identified using the 0.05 and 0.95 quantiles across 100 individuals, with values outside this range set to NaN. For quadratic analysis, data were smoothed by gestational week, and averages were computed. Data preprocessing involved calculating the number of days relative to each participant’s delivery date (days since delivery) at every signal and sample points, and determining the corresponding gestational age. Gestational age was calculated based on the estimated due date provided during the initial enrollment and adjusted according to actual birth date when necessary. This information was crucial in aligning wearable device data and survey responses with the appropriate stage of pregnancy for accurate longitudinal analysis. Fatigue was kept in its continuous form between 1-7 values where 1 is “none” and 7 is “severe”.

A case study analysis examined factors contributing to heterogeneity and varying signal behaviour among participants, including self-reported severe daily events, severe symptom events, and adverse event reports. Severe symptoms, outlined in Table S2, were reported bi-weekly. To enhance the evaluation of their impact on distribution change analysis, we applied the SentenceTransformer (26) model to generate embeddings for daily and bi-weekly features, enabling a more precise estimation of severe symptom timestamps.

Expanding this analysis, we examined the impact of adverse events on feature trajectories using a dataset subset with free-text descriptions recorded by engagement experts after bi-weekly check-ins. To ensure consistency and extract insights, we used ChatGPT to parse and label the free-text data, identifying explicit dates (MM/DD/YYYY or YYYY/MM/DD) and inferring incident dates from relative time references (e.g., ‘one week ago’). ChatGPT was prompted to generate concise labels for adverse events by recognizing key occurrences, like medical procedures or symptoms, and condensing them into meaningful terms. For example, ‘She started to get a cold 2 days ago, sinus and ear pressure, nasal congestion, and cough’ was labeled as ‘Infection,’ while ‘Participant went to the emergency room with body aches and a fever’ was labeled ‘Fever & Body Aches.’ This approach enabled efficient and consistent categorization of adverse events, ensuring high-quality labeled data for analysis. Further details, including the complete list of event labels and report counts, are provided in Table S5.

### Statistical Analysis

To assess the model’s performance, we calculated the coefficient of determination (R^2^), which quantifies how well the fitted quadratic model explains the variance in the observed data. An R^2^ value closer to 1 indicates a better fit and was estimated as:

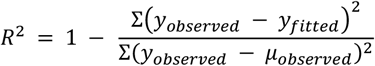

where y_observed_, μ_observed_, y_fitted_ represent the initial feature pattern, mean of observed data, and quadratic model output, respectively.

Kernel density estimation (KDE) is a non-parametric method for estimating the probability density function (PDF) of a continuous variable, providing a smooth data distribution representation. KDE places a Gaussian kernel on each data point and sums them to generate a continuous density estimate, avoiding parametric assumptions. To compare PDF distributions between subgroups, we applied the Kolmogorov-Smirnov test (27). A p-value < 0.05 indicates a statistically significant difference, rejecting the null hypothesis.

To evaluate the impact of including age as a predictor, we performed a likelihood ratio test comparing the full model with age to a reduced model without it. The log-likelihoods of both models were computed, denoted as ll_full_ for the full model and ll_reduced_ for the reduced model. The likelihood ratio statistic (LR statistic) was calculated as:

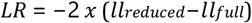

The degrees of freedom (df) for the chi-square distribution were determined by the difference in the number of parameters between the full and reduced models. Using the chi-square distribution, we computed the p-value (28) based on the LR statistic and df, providing statistical evidence for comparing the two models.

**For the case study analysis**, we first conducted an intra-individual assessment. For each event, we compared signal behavior around the event time (7 days before and after) to a baseline period (one month to 7 days prior). Comparisons were included only if the pre-event and around-event periods contained at least 7 data points, ensuring sufficient data for meaningful statistical analysis. We applied an unpaired Welch’s t-test, which does not assume equal variance, to evaluate the null hypothesis of equal means between the two periods. This test provided a t-statistic and p-value, indicating the statistical significance and direction of any differences in signal behavior. We then performed an inter-individual analysis to assess whether similar impacts were observed across individuals. Using 14-day windows from -270 to 0 gestational age, we examined the occurrence of each event within each window and calculated the average t-statistic for each individual. A high variability threshold was defined as twice the standard deviation of the average t-statistic across individuals. If the absolute difference in t-statistics between any two individuals exceeded this threshold, the event was flagged as having a distinct impact on feature patterns between individuals. For Figure 5c, participants and events were randomly selected from those meeting statistical significance criteria to ensure representativeness and minimize selection bias, illustrating examples of differing physiological responses across individuals.

## Results

### Aggregated analysis

The generalizability of aggregated results from three recently wearable studies (15–17) in maternal health was assessed by replicating and comparing them using a larger external wearable dataset, the BUMP study (19). These recently published papers examined: 1) HRV (15), 2) Fatigue (16), and 3) Sleep (17) aggregate patterns over time. The main findings included the following: Jasinski et al. (15) identified an inflection point in HRV around week 7 before birth for both term and preterm pregnancies, with an additional inflection at gestational week 33 for term pregnancies; an absence of this trend was noted in preterm cases; Nissen et al. (16) reported fatigue peaking early at gestational weeks 7–8 and reaching a trough around week 21; and Guo et al. (17) analyzed aggregate sleep physiology, noting a negative trend in deep sleep and a positive trend in awake time. Guo et al. also found that specific BMI and maternal age groups influenced these patterns, potentially helping in the identification of risk factors for adverse outcomes.

In the BUMP study, the aggregated results suggest that nighttime daily HRV shows a similar inflection point around week 7 before birth in both term and preterm pregnancies and at gestational week 33 in term pregnancies, though this trend is absent in preterm pregnancies (Figure 1a). However, the weekly mean HRV values differ significantly, with the BUMP data showing a smaller weekly variance compared to Jasinski et al. For fatigue, the general trends align between Nissen et al. ‘s paper and the BUMP study, except for the third trimester, with both showing an early peak around gestational weeks 7-8 and a trough around week 21 (Figure 1b).

**Figure 1.**
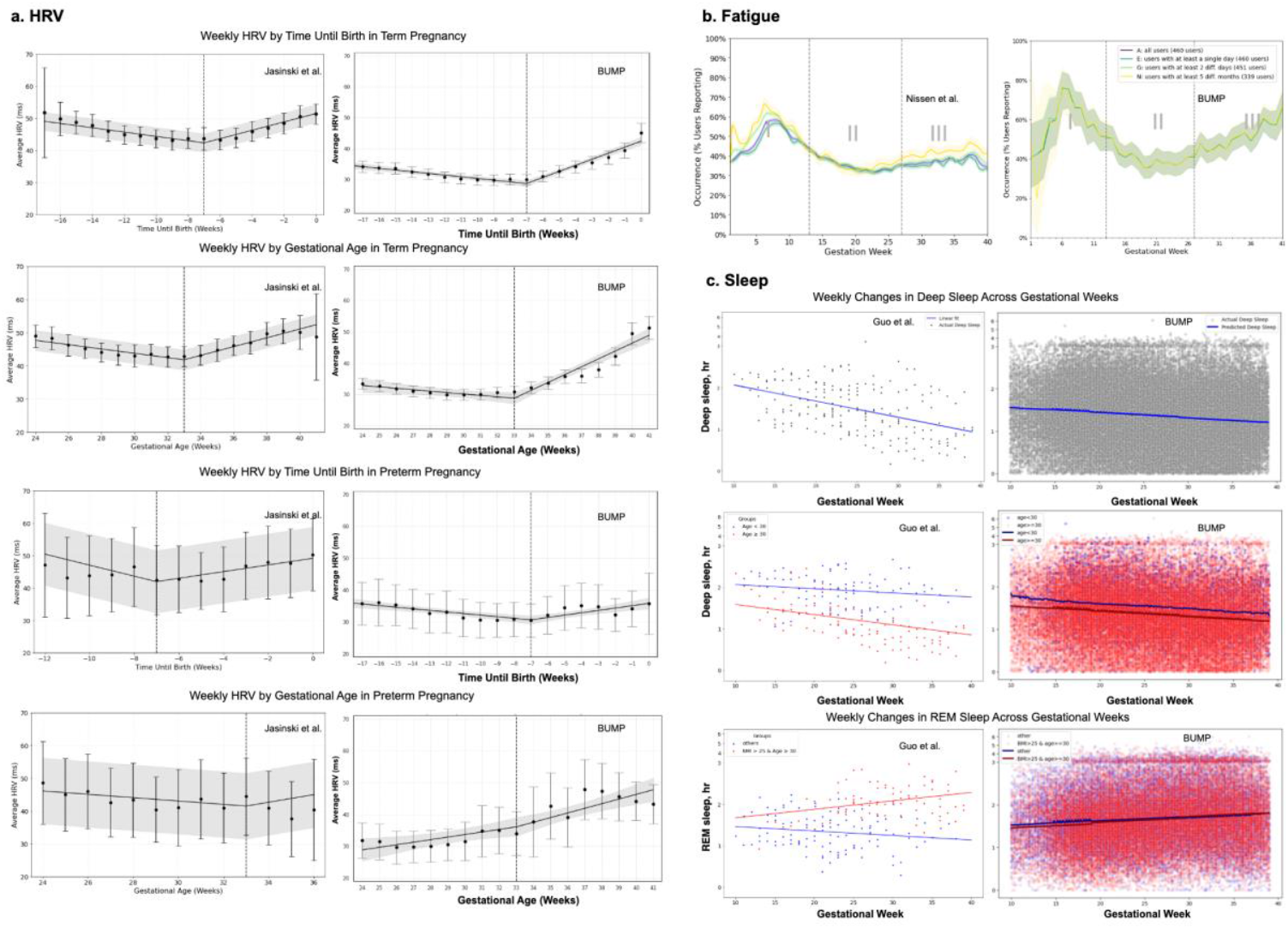
Aggregated Results Comparison of HRV, Fatigue, and Sleep Patterns Between BUMP (19) and Prior Pregnancy Studies. a)(15) HRV trends show similar pre-birth inflections, with less variance in BUMP. Error bars represent the 95% CI; the vertical dashed line marks the inflection point. b)(16) Fatigue aggregate peaks and troughs pattern align with Bump. Dashed vertical lines mark the pregnancy trimesters. c)(17) Deep sleep trends differ in slope with less variation across BMI and age subgroups in BUMP.

Regarding deep sleep (Figure 1c), both the BUMP data and Guo et al.’s findings show a negative slope for aggregate deep sleep, with Guo et al. reporting a slightly steeper decline across 10–40 gestational weeks. Our analysis further reveals substantial variability and heterogeneity in sleep data, reflected in Intraclass Correlation Coefficients (ICCs) of 0.618 for deep sleep and 0.502 for REM sleep, both exceeding the 0.5 threshold for significance. In the BUMP data, BMI and age were not clearly separable predictors, unlike Guo et al.’s findings. Segmenting age into ≥30 years vs. <30 years did not significantly improve model fit (p=0.17), suggesting that Guo et al.’s proposed associations may not be generalizable to all populations. Similarly, stratifying the data by pre-pregnancy BMI and age (age ≥30 & BMI >25 vs. others) yielded a p-value of 1, further questioning the generalizability of these demographic factors as predictive indicators.

These findings highlight that results from wearable maternal health studies are not easily generalizable across populations due to significant heterogeneity in wearable signal patterns and individual subjective symptom experiences.

The analysis of HRV, fatigue, and sleep metrics derived from personal DHT data during pregnancy underscores the limitations of aggregated models in capturing individual variability in maternal health. Figure 2 shows violin plots, using density curves to illustrate data distribution. Wider sections indicate higher data density, enabling intuitive comparisons of individual variability against aggregate trends. Grey lines mark key findings from each study, contrasted with BUMP dataset analyses to assess density across individual-level data.

**Figure 2.**
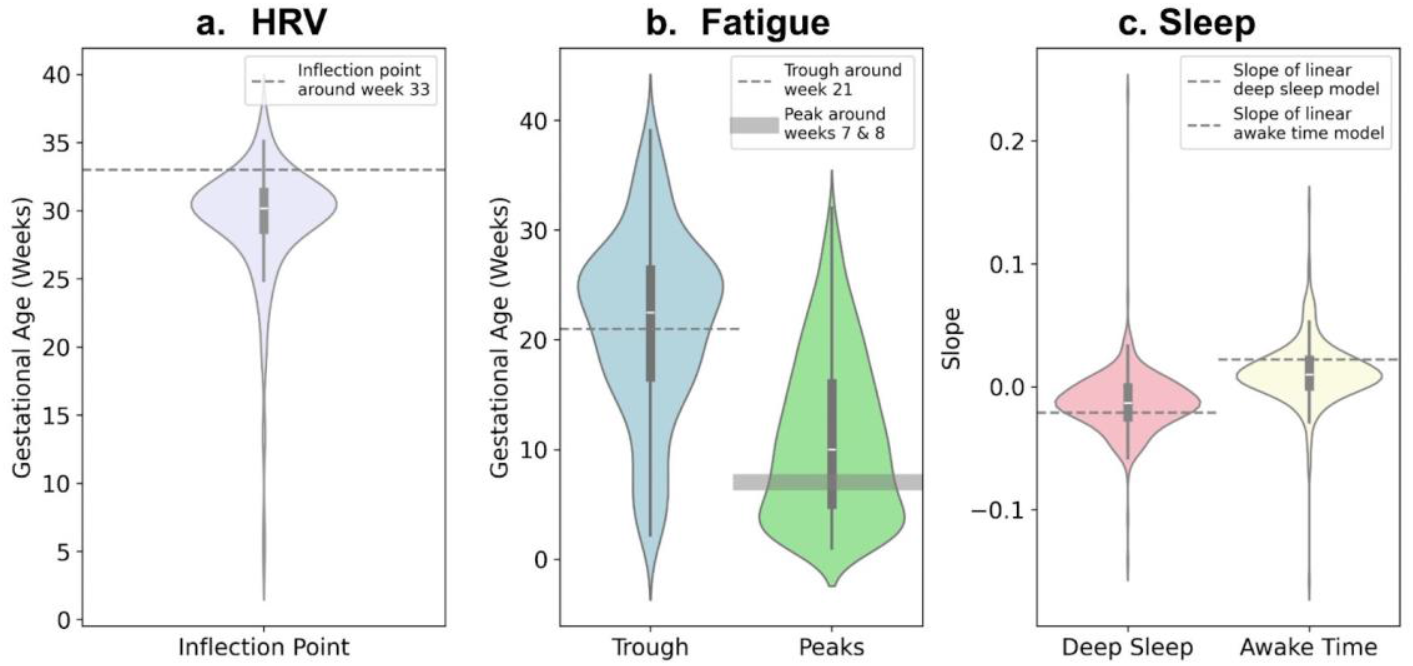
Key Features Extracted from Wearable Data During Pregnancy in the BUMP Study. Horizontal hashed or highlighted lines represent the findings from Jasinski et al, Nissen et al, and Guo et al based on aggregate results. The violin plots compare these aggregate findings with individual-level BUMP analysis, highlighting variability across participants.

Key comparisons include the following: For HRV, Jasinski et al. identified an inflection point around gestational week 33. To compare with individual-level inflection points, BUMP data was analyzed using a quadratic model (weeks 10–40), identifying the minimum point as the individual inflection. Similarly, aggregate fatigue trends in BUMP aligned with Nissen et al., showing early peaks at weeks 7–8 and a trough near week 21. Individual fatigue peaks and troughs were calculated to capture variability. For sleep, Guo et al. used a linear mixture model to estimate slopes for aggregated deep sleep and awake time, which were compared with individual-level slopes determined by fitting linear regression models.

Figure 2 and Table S7 show that individual-level results from BUMP, visualized through density curves, did not fully align with the published findings and revealed substantial variation. Figure 2b highlights that individual fatigue troughs ranged from weeks 8 to 38, with a median at week 23; two weeks earlier than the aggregate result. Similarly, individual fatigue peaks occurred two weeks later than the aggregate finding, with notable variability indicated by the interquartile range and density spread. These discrepancies underscore the need to account for individual variability, as aggregate findings do not universally apply, even to the majority of individuals.

### N-of-1 level analysis

Individual patterns in HRV, fatigue, deep sleep, and awake time were examined and compared to aggregate trends using BUMP study data (Figure 3).

**Figure 3.**
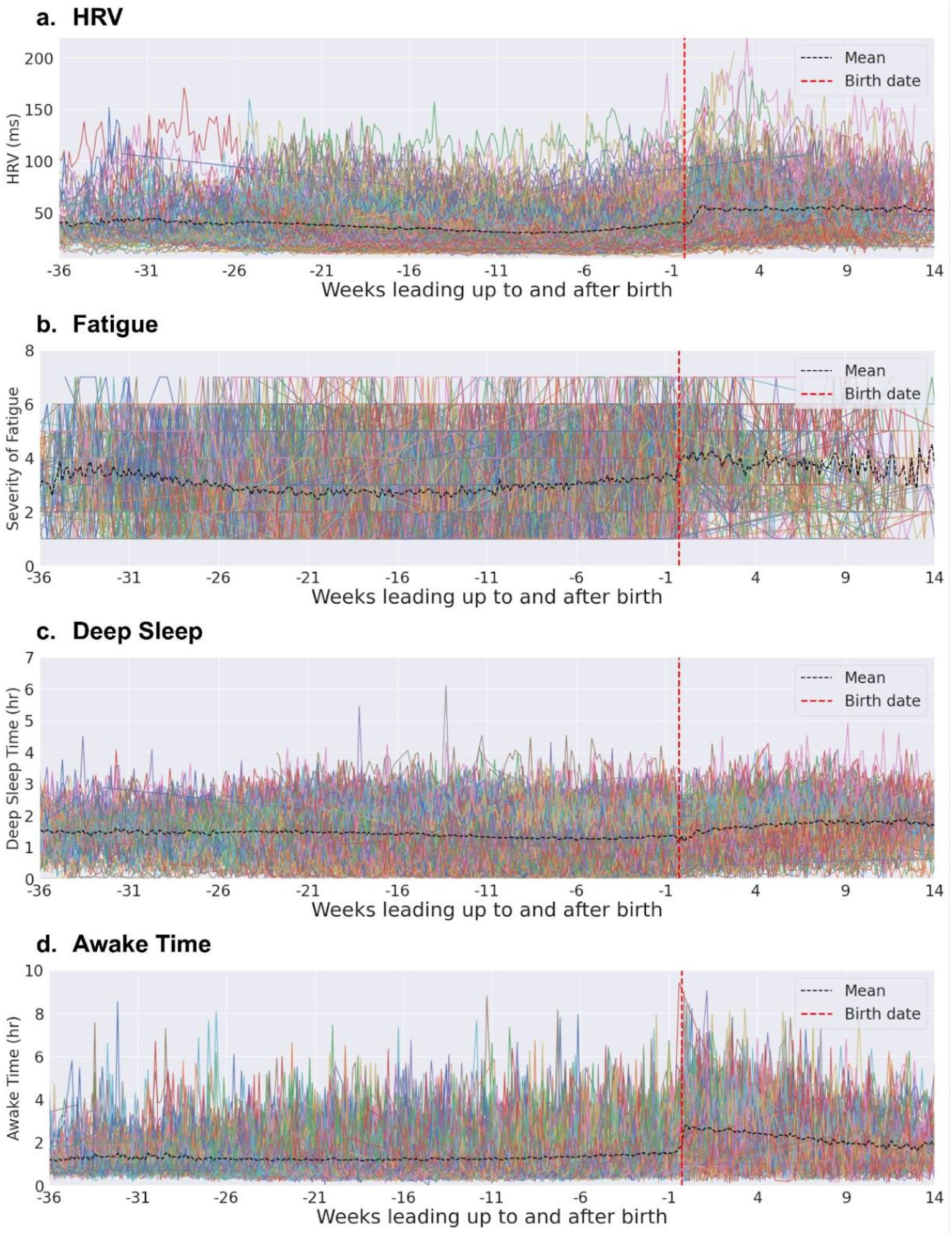
Spaghetti plots of four features from the BUMP study contrast aggregate trends with individual trajectories. Each colored line represents an individual’s HRV, fatigue, awake time, or deep sleep pattern, highlighting variability across gestational weeks.

Figure 3 highlights the high variability in objective and subjective feature patterns during pregnancy, often lost in aggregate views. Individual HRV, fatigue, deep sleep, and awake time patterns vary significantly across gestational weeks. While the aggregate line (black) suggests a smooth trend, it overlooks individual complexities and key variations. Relying solely on aggregate data risks misinterpretation, especially when individual patterns deviate from the average.

Figure 4 presents kernel density plots and p-values for R^2^ values assessing model fit across pregnancy-related complications. Modifying factors or confounders such as pregnancy-related complications and demographic factors may in part explain the observed heterogeneity in the model’s performance. However, the kernel density plots and p-values revealed no significant differences in R^2^ distributions, indicating that the model’s fit is relatively consistent across groups. Specifically, there were no significant differences in HRV, fatigue, deep sleep, or awake time trends among participants with different pregnancy-related conditions. All subgroup comparisons yielded p-values >0.05 (see lower section of Figure 4). The symmetric, overlapping kernel density plots suggest minimal variation in R^2^ distributions, with no skewness or distinct clustering. This analysis highlights that pregnancy-related complications and demographic factors (as shown in Figure S1) cannot be meaningfully separated into distinct subgroups, as these factors do not significantly impact the model’s ability to fit outcomes.

**Figure 4.**
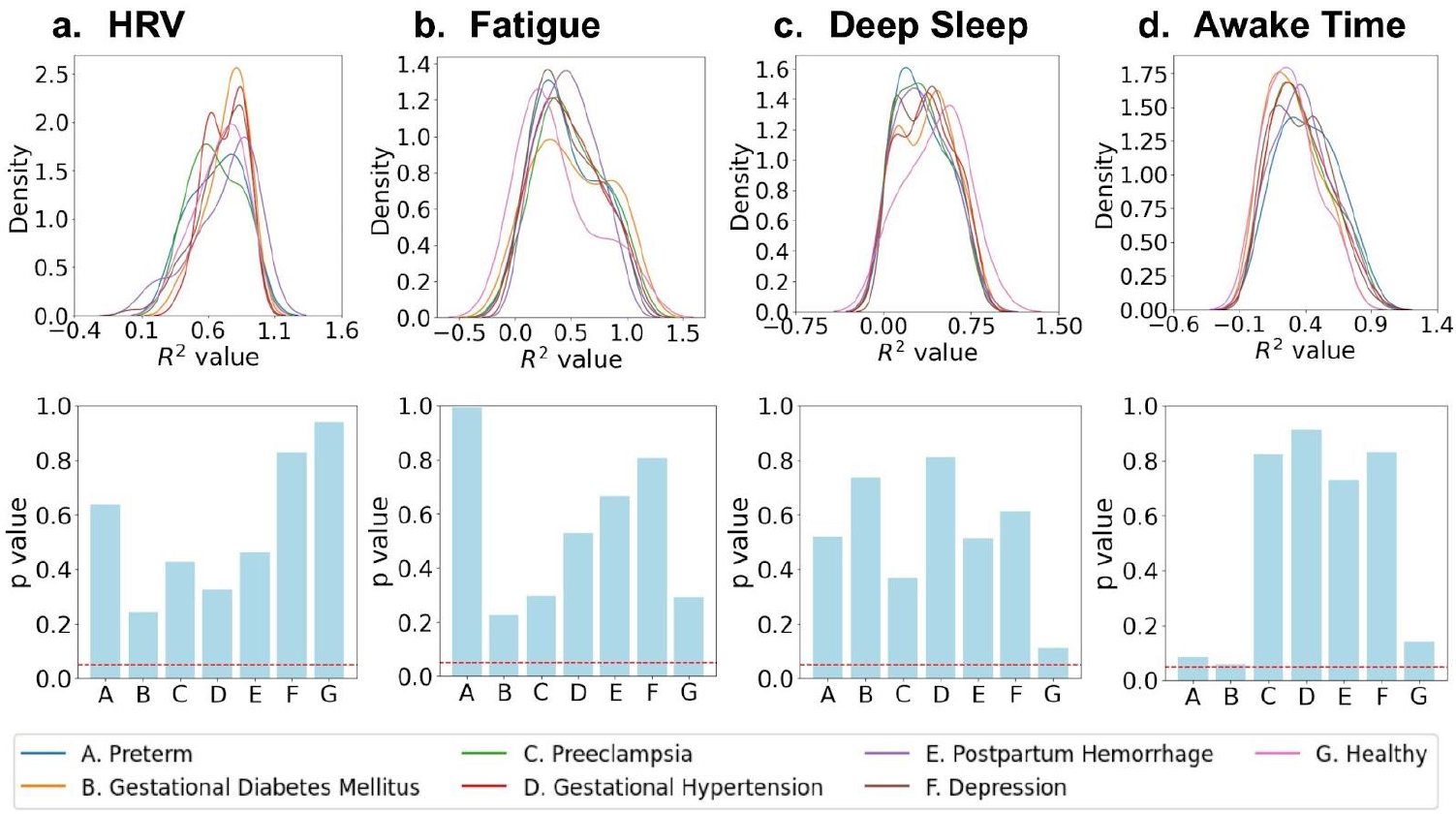
Comparison of R^2^ values assessing model fit across pregnancy-related complications in the BUMP study. Top plots illustrate kernel density distributions for each group, while bottom plots display p-values, comparing each group’s feature distribution to that of other women. The red dashed line marks the significance threshold (p = 0.05). Kernel density plots and p-values indicate no significant differences in R^2^ distributions between complications, suggesting the model’s fit remains consistent across groups.

### Case Studies

To explore sources of heterogeneity, we analyzed survey data on severe symptoms reported by users and adverse events (AEs) documented in research staff notes. Key events and their precise timing were identified from unstructured AE notes using GPT-based automatic prompting. We also incorporated daily severe symptoms: very low energy, impaired cognition, very low mood, and extreme stress, referred to as Severe Daily Features. Biweekly severe symptom surveys, aligned with daily reports, covered severe fever or cough, intense headaches, severe infections, vision changes, and urinary issues. Further details are available in the Methods section and supplementary material.

Figure 5a presents examples of high-frequency adverse events and severe symptoms reported during pregnancy. Among 257 individuals, 4,674 events were recorded, with a median of 9 and an average of 21 events per person, ranging between 1 to 153. Figure 5b highlights two participants randomly selected for significant shifts in feature distributions (p < 0.05) before and after severe events. In participant C, a low-energy event 155 days before delivery led to increased fatigue, awake time, and decreased HRV and deep sleep. In participant D, a severe infection 92 days before delivery had similar effects.

**Figure 5.**
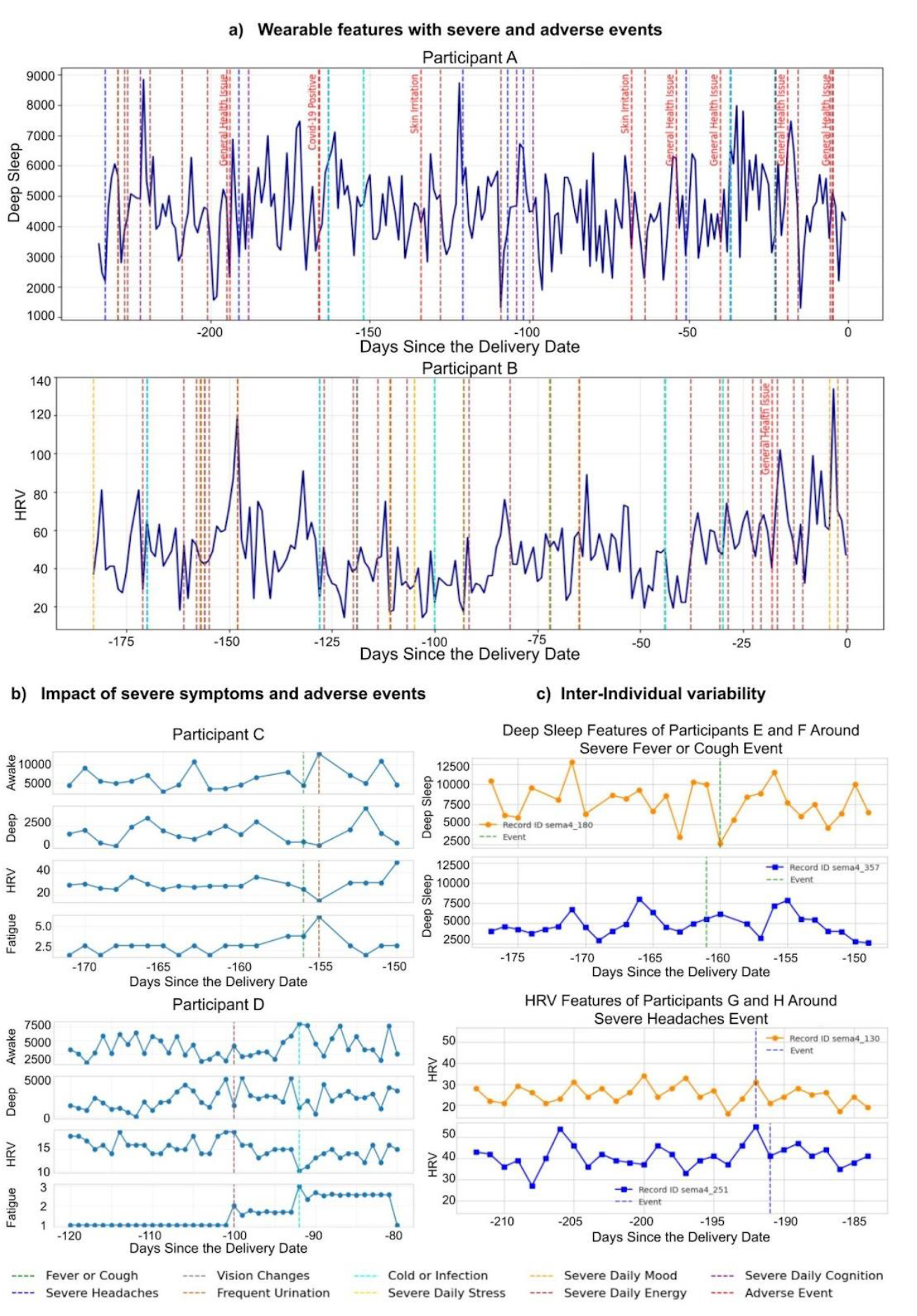
Impact of adverse events and severe symptoms on fatigue, HRV, and sleep features over pregnancy. a) Effects of adverse or severe events on deep sleep and HRV in example BUMP participants, with adverse events shown in red next to hashed lines. b) Examples of adverse or severe events significantly impacting the distribution of features throughout pregnancy. c) Despite experiencing the same severe symptom at the same gestational week, two women showed different sleep and HRV patterns.

Figure 5c illustrates the varying effects of severe symptoms on pregnant women at the same gestational week. Participants were randomly selected from those with significant physiological responses to the same symptoms, highlighting individual variability in adaptation. For example, participants E and F both experienced severe fever, sore throat, and/or cough in the same week, yet one had a sharp decline in deep sleep while the other showed a gradual increase. A linear mixed model analysis revealed substantial group-level variability for all events. These findings underscore the need to consider both inter- and intra-individual differences when evaluating responses to similar events. Additional examples are shown in Figure S4.

## Discussion

There is significant heterogeneity of self-reported symptoms and objective physiologic features over pregnancy. While some health measures follow average trends, these patterns often misrepresent many pregnant individual experiences. This highlights a significant limitation of aggregated models in maternal health, as they often fail to capture the complexity and variability of individual trajectories. A major advantage of DHTs is their ability to track individualized health trajectories. However, most current applications rely on aggregate analytics, limiting the full potential of this rich data for personalized monitoring and oversimplifying the diverse physiological and subjective changes. Here, we replicated aggregate analyses from three recent maternal health studies examining HRV (15), sleep (17) and self-reported fatigue (16) during pregnancy using an external maternal health dataset (BUMP).

The aggregate results from these studies, and from the BUMP dataset demonstrate unique contours in these three measures of health. Notably, HRV appears to steadily decrease with gestational age, but then shows an inflection at approximately 33 weeks leading up to delivery. This inflection has been highlighted in the literature by others, suggesting a phenomenon that could reflect an individual’s physiological readiness for delivery (15,21,22). While this could be true for some, our N-of-1 analyses demonstrate that many individuals do not experience this HRV inflection, or an inflection is experienced, but in a significantly different pattern from the average. Fatigue aggregate trends show an early peak around gestational weeks 7-8, followed by a gradual decline to a trough near week 21 (16). However, individual analyses reveal considerable variation in these patterns, as shown in Figure 2. Similarly, while average sleep trends show a steady decline in deep sleep and increased awake time, especially in the third trimester (17,23), N-of-1 analyses highlight substantial inter-individual differences. Some individuals experience a progressive decline in deep sleep, while others show minimal or even opposite changes, underscoring the complexity of sleep adaptations during pregnancy.

Physiological and subjective changes during pregnancy often differ from group averages due to external factors. While previous studies (5,17) have often relied on standard demographic exclusions or subgrouping and stratified analyses by age or pregnancy complications, these approaches miss the substantial heterogeneity in individual patterns. Using N-of-1 analyses with spline models, we captured each participant’s unique response to pregnancy-related changes, offering detailed insights into individual trajectories. Our N-of-1 analyses found no significant differences in HRV, sleep and fatigue levels between participants with pregnancy complications and those without, suggesting these conditions do not fully explain the observed heterogeneity. Some differences emerged across demographic factors like age and BMI; for instance, individuals with a BMI of 25–30 showed significant differences in HRV and deep sleep goodness-of-fit compared to other subgroups. However, this pattern wasn’t observed for fatigue or awake time, indicating BMI alone isn’t a reliable indicator for subgrouping but warrants further exploration alongside other factors.

We further explored case studies in an attempt to isolate potential modifying factors of the examined maternal health metrics. The case studies further compounded the finding suggesting high heterogeneity of maternal health experiences. For example, while in some participants there were associations between events such as fever, cough, and very low energy levels and significant increases in fatigue and awake time, there was high intra- and inter-individual variability in responses to these events emphasizing that pregnant individuals can exhibit diverse physiological responses to the same event at the same gestational week.

Taken together, after extensive exploration into potential modifiers of HRV, sleep, and fatigue over pregnancy, we identify no clear factors explaining the observed high heterogeneity of these maternal health metrics, suggesting the necessity of personalized analytical approaches to accurately interpret maternal health data at N-of-1 level. The significant variability among individuals highlights the complexity of deriving broadly applicable clinical insights.

In conclusion, this study underscores the need for personalized and diverse assessments to capture the individuality of maternal health metrics. Moving beyond one-size-fits-all models is essential for embracing heterogeneity and tailoring insights to pregnant individuals’ unique needs. Future work should prioritize developing personalized methods that account for variability and assess causal effects of modifying factors. Effectively interpreting personal DHT maternal health data may require human-in-the-loop approaches, where users are prompted for additional context when deviations in signals are detected.

## Acknowledgments

This study was funded by 4YouandMe and Sema4, with additional in-kind contributions from coalition partners including Evidation Health, Vector Institute, Cambridge Cognition, and Bodyport. Additional support was provided by the Varma Family Chair (AG) and CIFAR AI Chair (AG). We would also like to thank all the women who participated in the BUMP and BUMP-C studies for their ongoing commitment and valuable contributions to enhancing future pregnancy care.

## Funding Statement

This work was supported by 4YouandMe, a nonprofit organization based in Seattle, WA, USA.

## Conflicts of Interest Statement

All authors declare no financial or non-financial competing interests.

## Data and Code Availability

Among participants who opt-in, coded study data from the BUMP study participants is available on the Synapse platform (synapse.org) at Sage Bionetworks (https://sagebionetworks.org) and can be freely accessed by any researcher who becomes ‘qualified’ by becoming a registered and certified Synapse user (https://help.synapse.org/docs/User-Account-Tiers.2007072795.html), and by meeting the specific conditions of use that require submitting an intended data use statement alongside an IRB approved protocol. The BUMP specific Synapse Project page can be found here: https://www.synapse.org/#!Synapse:syn25953345/wiki/616547 among registered Synapse users.

The BUMP study was ethically approved by the Institutional Review Board, Advarra (Pro00047893). E-consent was obtained from all participants in the study app.

Reproducibility Github link and website page: https://github.com/Tinbeh97/N-of-1-Level-Wearable-Data-Analysis

https://tinbeh97.github.io/N-of-1-Level-Wearable-Data-Analysis/?token=n1wearable123

## Author Contributions

JY and TB designed the methodology, conducted the formal analysis, developed the software, interpreted and validated the results, and drafted the original manuscript. RY contributed to methodology design, analysis, software development, result interpretation, validation,data curation, and manuscript review and editing. AB assisted with data curation, software development, analysis, and manuscript review and editing. AG and SG contributed to conceptualization, supervised the analysis, interpreted results, and reviewed and edited the manuscript. SF led the overall study design and conceptualization, secured funding, supervised the analysis, interpreted results, and reviewed and edited the manuscript. All authors have read and approved the final manuscript. TB, JY contributed equally to this manuscript.

## List of Abbreviations

DHT: Digital Health Technologies
HRV: Heart Rate Variability
AE: Adverse Events
GDM: Gestational Diabetes Mellitus
ICC: Intraclass Correlation Coefficient
BUMP study: Better Understanding the Metamorphosis of Pregnancy study
RMSSD: Root Mean Square of Successive Differences Between Normal Heartbeats
BMI: Body Mass Index
REM: Rapid Eye Movement

## Notes

### Competing Interest Statement

The authors have declared no competing interest.

### Funding Statement

The study was funded by 4YouandMe(nonprofit), Seattle, WA, USA.

### Author Declarations

The BUMP study was ethically approved by the Institutional Review Board/IRB, Advarra (Pro00047893).

