## Supplementary Material for "Maternal health Aggregated Trends can be Misleading: The power of N-of-1 Level Wearable Data Analysis for Personalized Pregnancy Monitoring"

**This PDF file includes:**

Supplemental Information Text
Supplemental Figures 1 to 4
Supplemental Tables 1 to 7
Supplemental References

### Supplementary Methodology

- **Comparison Section Methodologies**

A linear mixed model (LMM) was employed in this study to account for both fixed and random effects, allowing us to model individual variability while examining the relationship between predictor variables and outcomes of interest. The fixed effects represent the overall population-level impact of variables such as gestational age, demographic factors, and physiological metrics, while the random effects capture the subject-specific deviations from the population mean. The model was fitted using maximum likelihood estimation, and key performance metrics, such as the log-likelihood and Intraclass Correlation Coefficient (ICC), were used to evaluate model fit and variability between individuals.

- **Data Collection and Study Measures**

The Oura Ring gen 2 and 3 was used to capture features such as sleep duration, heart rate variability, and activity levels. The HRV, deep sleep, awake time, and fatigue measures were selected based on their impact on pregnancy and data availability and to explore n-of-1 level analyses for maternal health from multiple perspectives. HRV was calculated using beat-to-beat intervals derived from photoplethysmography (PPG) signals collected at a frequency of 50Hz, with robust artifact detection and filtering methods ensuring accuracy (1). HRV offers an important measure to analyze during pregnancy, as it provides a non-invasive measure of the maternal autonomic nervous system (2), potentially acting as a digital biomarker for preterm birth (3).

Sleep, which undergoes significant changes during pregnancy, is also critical for both maternal and fetal health (4). Sleep stages, including wake, light sleep, deep sleep, and REM sleep, were classified at 5-minutes intervals using multi-sensor data. Awake time was determined based on accelerometer readings, while deep sleep was identified using a combination of heart rate variability, body temperature, and accelerometer data collected via the Oura Ring (1). Both awake time and deep sleep were included in the analysis, as awake time is uncorrelated with HRV, whereas deep sleep patterns may exhibit some correlation (5).

A BUMP study smartphone app (6) was used to track daily, weekly, or bi-weekly self-reported surveys of pregnancy related symptoms and other socio-demographic factors. Pregnancy-related complications, including gestational hypertension, gestational diabetes, preeclampsia, eclampsia, toxemia, preterm birth, and postpartum depression, were assessed using a multi-source data approach. Participants completed a post-birth phone survey with a research coordinator between one and three months postpartum, and this data was supplemented with electronic health record (EHR) information obtained through Sema4's patient platform with participants' digital consent[19].

Participants were also prompted to complete a one-time demographic survey that collected information such as age, ethnicity, body mass index (BMI), and socioeconomic status. Additionally, participants completed a daily self-reported symptom survey to rate the severity of pregnancy-related symptoms. For this analysis, the fatigue item from the survey was used. Participants were asked, *"In the past day, have you noticed any symptoms? Feeling fatigued or easily tired?"* Responses were recorded on a scale from 1 to 7, with 1 indicating minimal fatigue and 7 indicating severe fatigue. Fatigue, a prevalent and subjective symptom, further broadens the understanding of maternal well-being by highlighting potential health concerns (7).

Additional self-reported information was captured over the phone with study engagement specialists on average every two weeks. Specifically, adverse events (AEs) were documented using a dedicated adverse event questionnaire, allowing for detailed tracking of any health complications or concerns during the study period. A post birth survey at approximately one-month postpartum was also conducted over the phone that asked participants questions about their delivery experience.

**Data exclusion:** We excluded individuals whose quadratic fits to deep sleep objective values had vertices outside the meaningful range (10–40 gestational weeks), as these likely reflected implausible patterns caused by noise or incorrect recordings, potentially compromising the validity of deep sleep measurements and introducing bias.

- **Generative AI full description**

The Model: ChatGPT-4-turbo, accessed through OpenAI's API
Tasks and Prompts:

1. Extracting incident dates from free-text descriptions

    a. "Look for any dates in the ae_describe column that are formatted as MM/DD/YYYY, YYYY-MM-DD, or MM/DD/YY and extract them into a new column called incident_date. If only Month/Day is provided, use the corresponding year from the date column. If the description includes relative time references like '1 week ago' or '3 days ago', calculate the incident_date by subtracting the specified time from the date column."

2. Generating Structured Labels from Free-Text Descriptions

    a. "Based on the ae_describe column, create a column called label *that* provides a short, meaningful keyword summarizing what happened. For example:

        i. 'COVID-19 infection' → 'Covid-19 Positive'

        ii. 'Participant went to the emergency room for body aches and fever' → 'Fever & Body Aches'

    b. Generate consistent labels across similar events, ensuring no generic terms like 'Other' are used."

3. Refining COVID-19 Labels Based on Description Context

"For labels that include 'Covid-19', ensure that the description in ae_describe confirms a positive COVID-19 test. If the description includes 'negative' or indicates the absence of COVID-19, remove the label. All other relevant cases should be uniformly labeled as 'Covid-19 Positive'."

**Supplementary Analysis**

Figure 1c further statistical analysis: The Bayesian Information Criterion (BIC) for the full model is higher than the reduced model, with a no change and 19.9% decrease for deep and REM sleeps, respectively, after model reduction. This indicates that adding demographic segmentation does not improve the detection of changes in deep or REM sleep.

Delivery information: Summary statistics for gestational weeks at the time of delivery: mean = 38.49, standard deviation = 2.07, minimum = 26, maximum = 41.

**Supporting Information Figures**

a. **HRV**

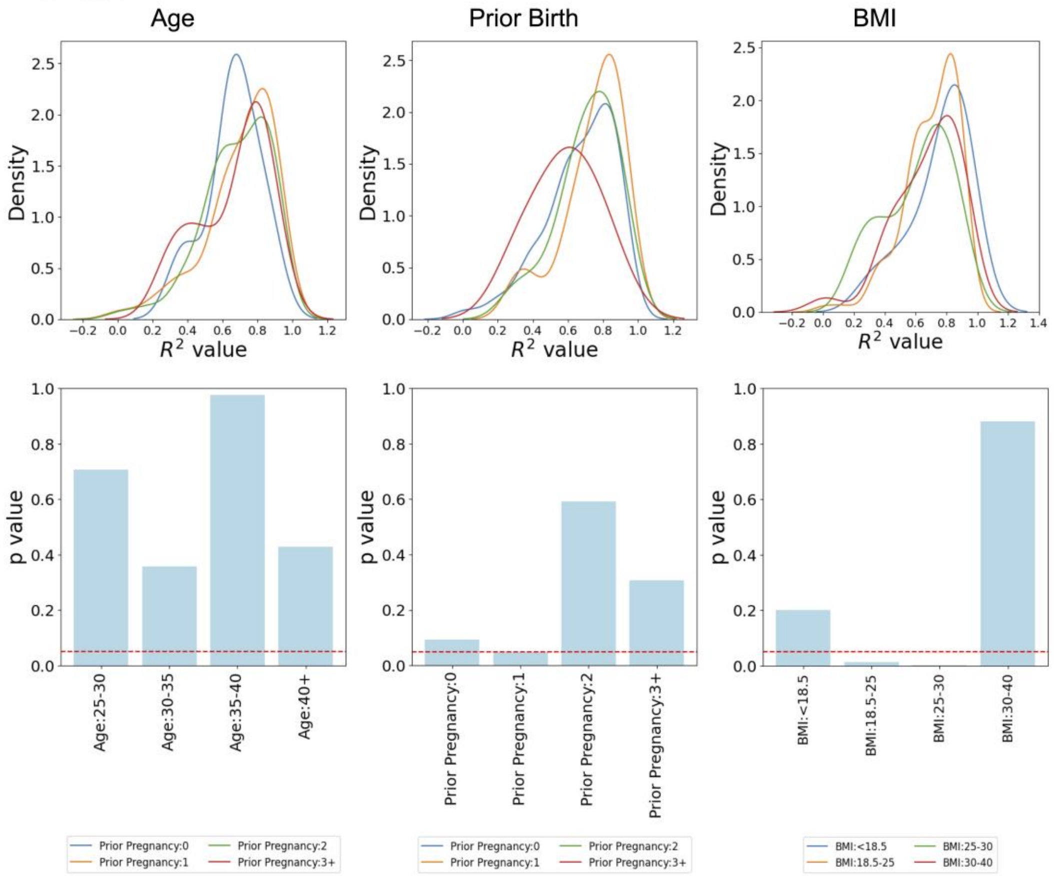

### b. Fatigue

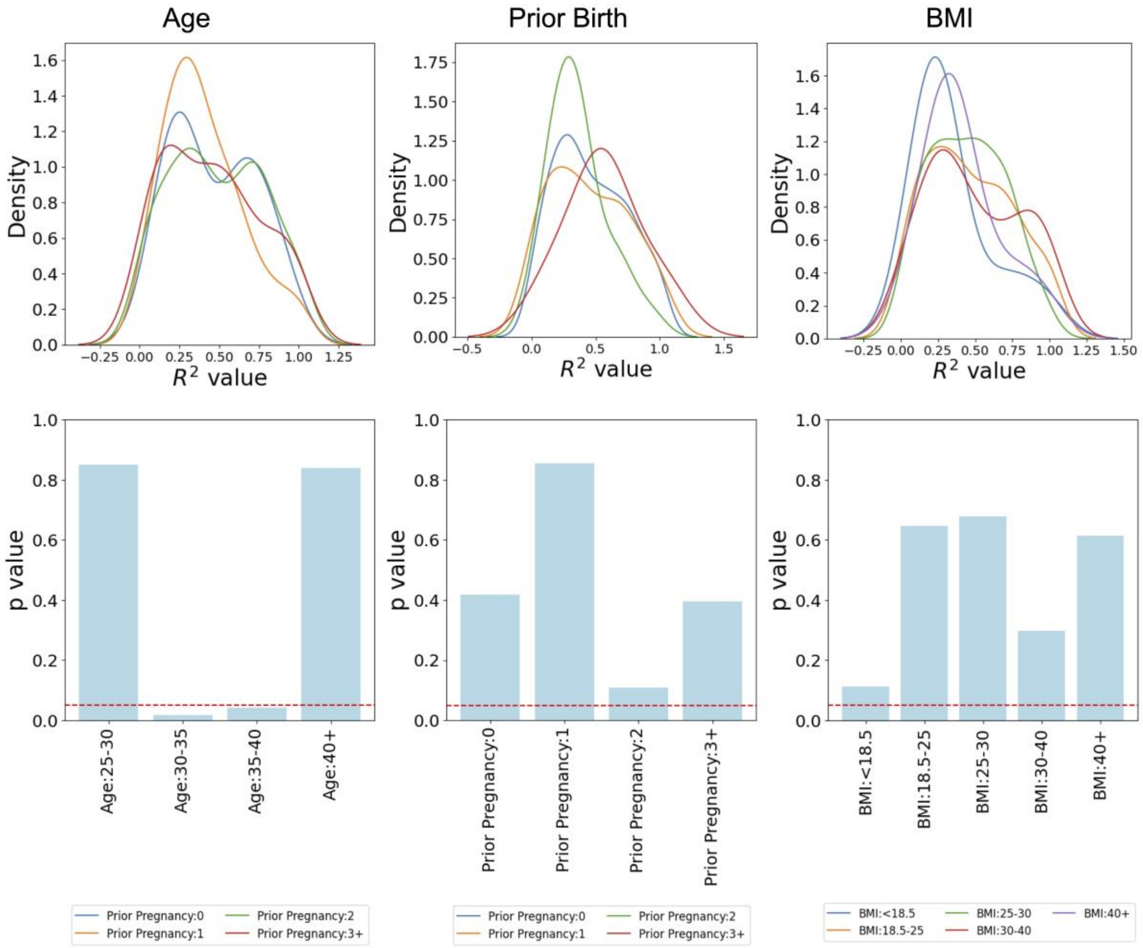

Age
Prior Birth
BMI
Density
$R^2$ value
p value
Age:25-30
Age:30-35
Age:35-40
Age:40+
Prior Pregnancy:0
Prior Pregnancy:1
Prior Pregnancy:2
Prior Pregnancy:3+
BMI:<18.5
BMI:18.5-25
BMI:25-30
BMI:30-40
BMI:40+

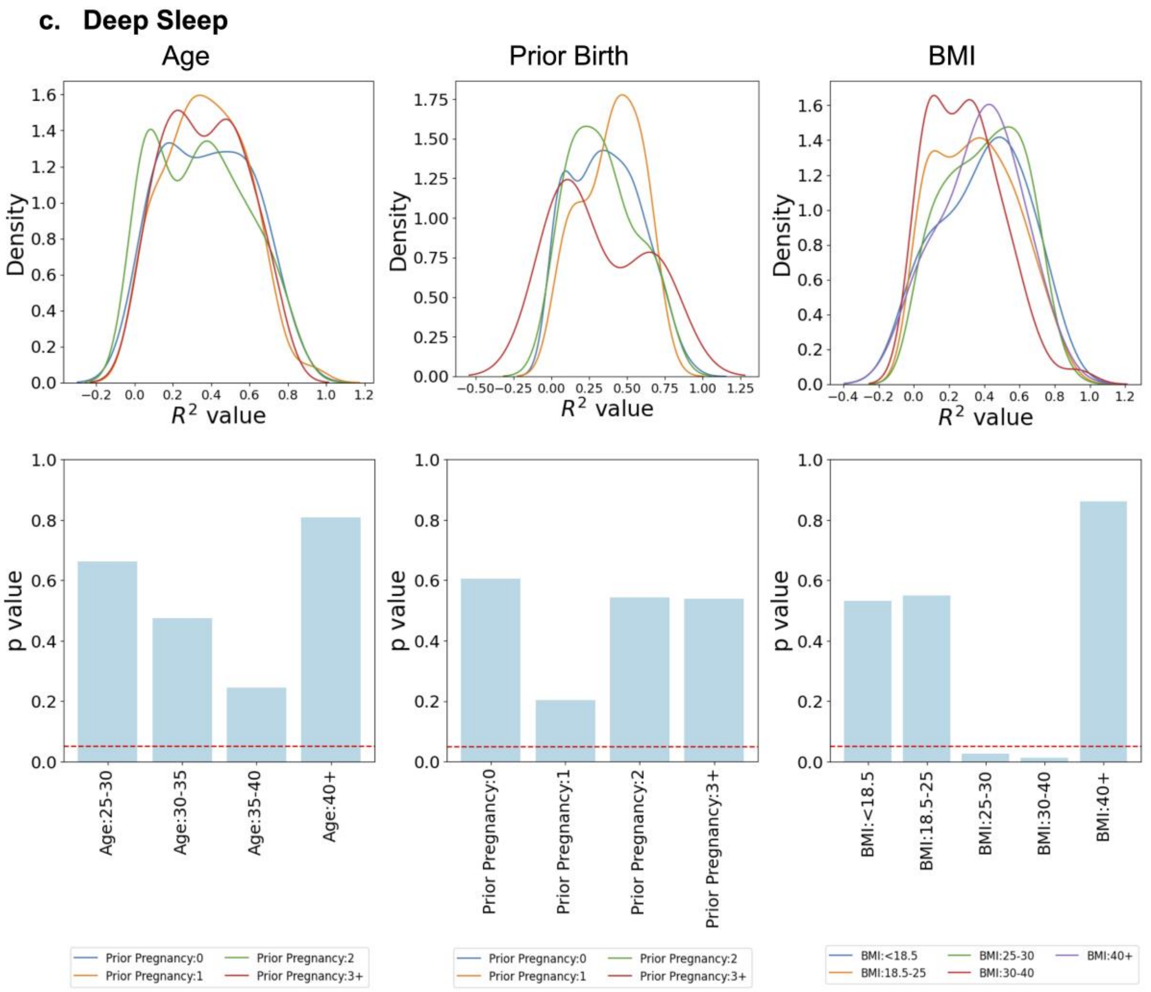

c. Deep Sleep
Age
Prior Birth
BMI
Density
$R^2$ value
p value
Age:25-30
Age:30-35
Age:35-40
Age:40+
Prior Pregnancy:0
Prior Pregnancy:1
Prior Pregnancy:2
Prior Pregnancy:3+
BMI:<18.5
BMI:18.5-25
BMI:25-30
BMI:30-40
BMI:40+

**d. Awake**

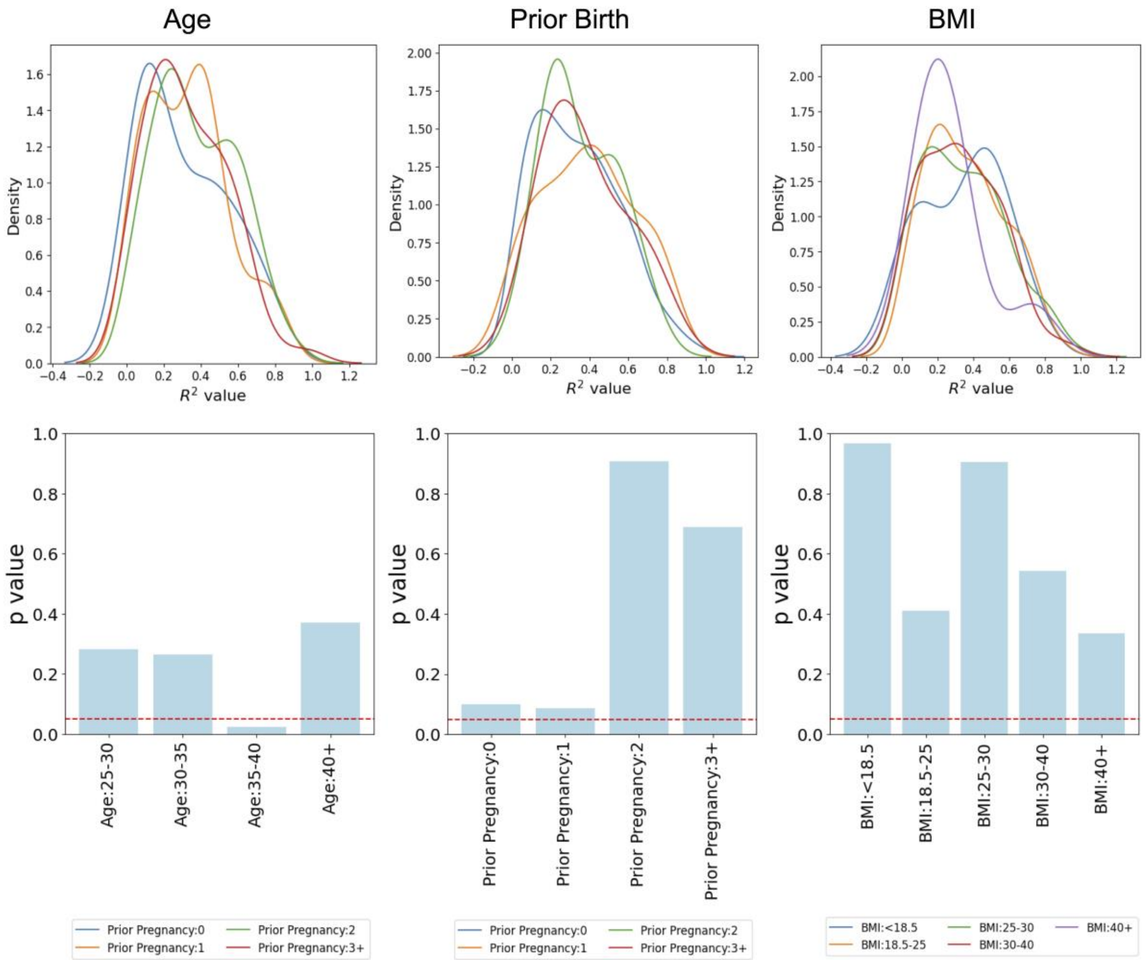


**Fig. S1. Comparison of $R^2$ values assessing goodness of fit across different age, BMI, and parity categories in women.**

The demographic analyses, including age, prior births, and BMI, were conducted across four key features: HRV, Fatigue, Deep Sleep, and Awake Time. Here, $R^2$ values from the spline model were compared to evaluate the goodness of fit for various pregnancy-related complications using data from the BUMP study.

The age cut-off values are categorized as 25–30, 30–35, 35–40, and 45+. BMI groups are defined as <18.5, 18.5–25, 25–30, 30–40, and 40+. Parity groups are classified as 0, 1, 2, and 3+ (more than 3 previous births). The 25–30 BMI group shows a significant difference in $R^2$ values ($p < 0.05$) for HRV and Deep Sleep, although the number of participants in this group is limited. Additionally, the 35–40 age group significantly impacts Awake Time and Fatigue $R^2$ goodness of fit, indicating that it is statistically different from other groups in model performance.

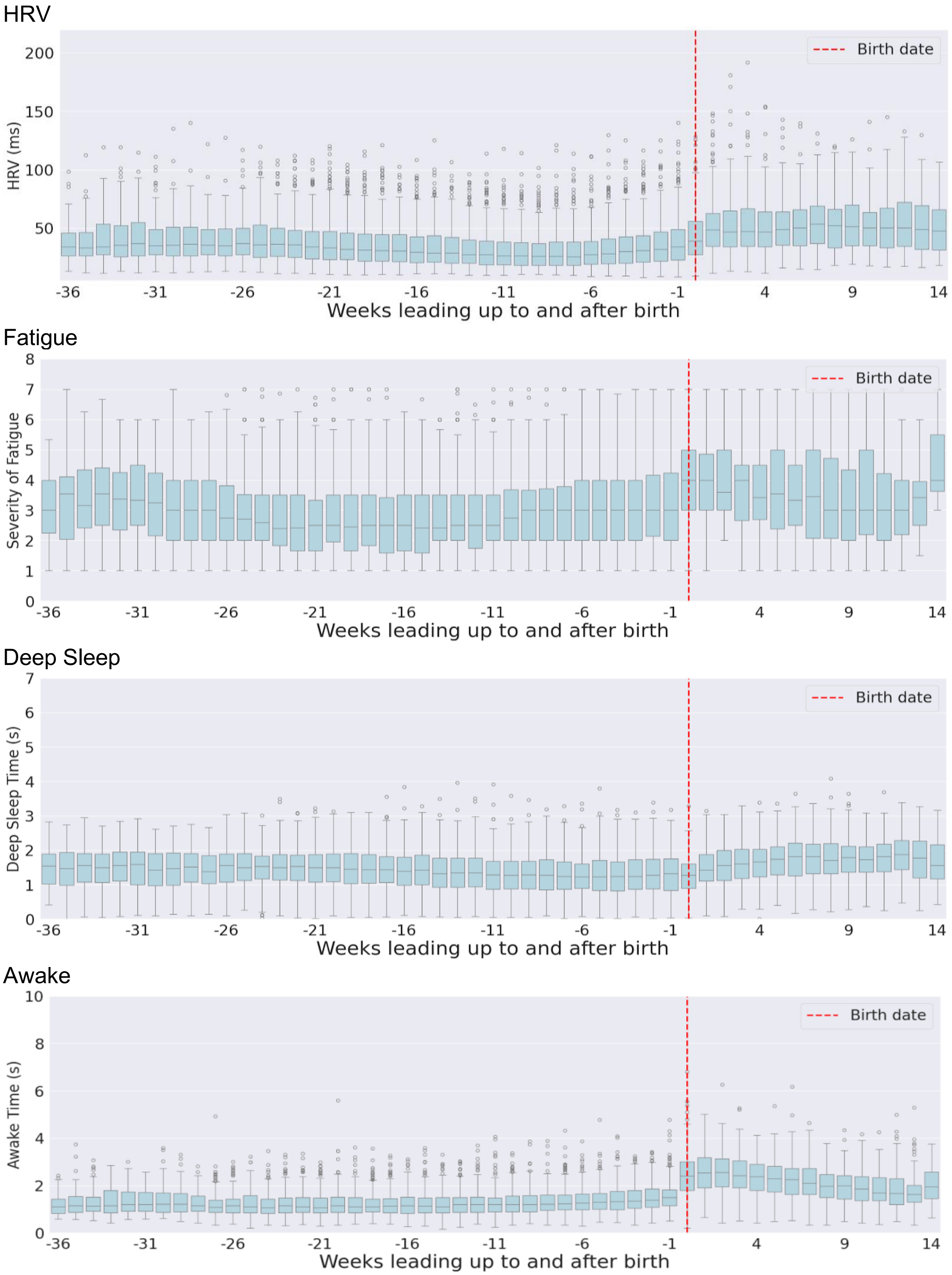


**Fig. S2. Box plots of four features during pregnancy.**

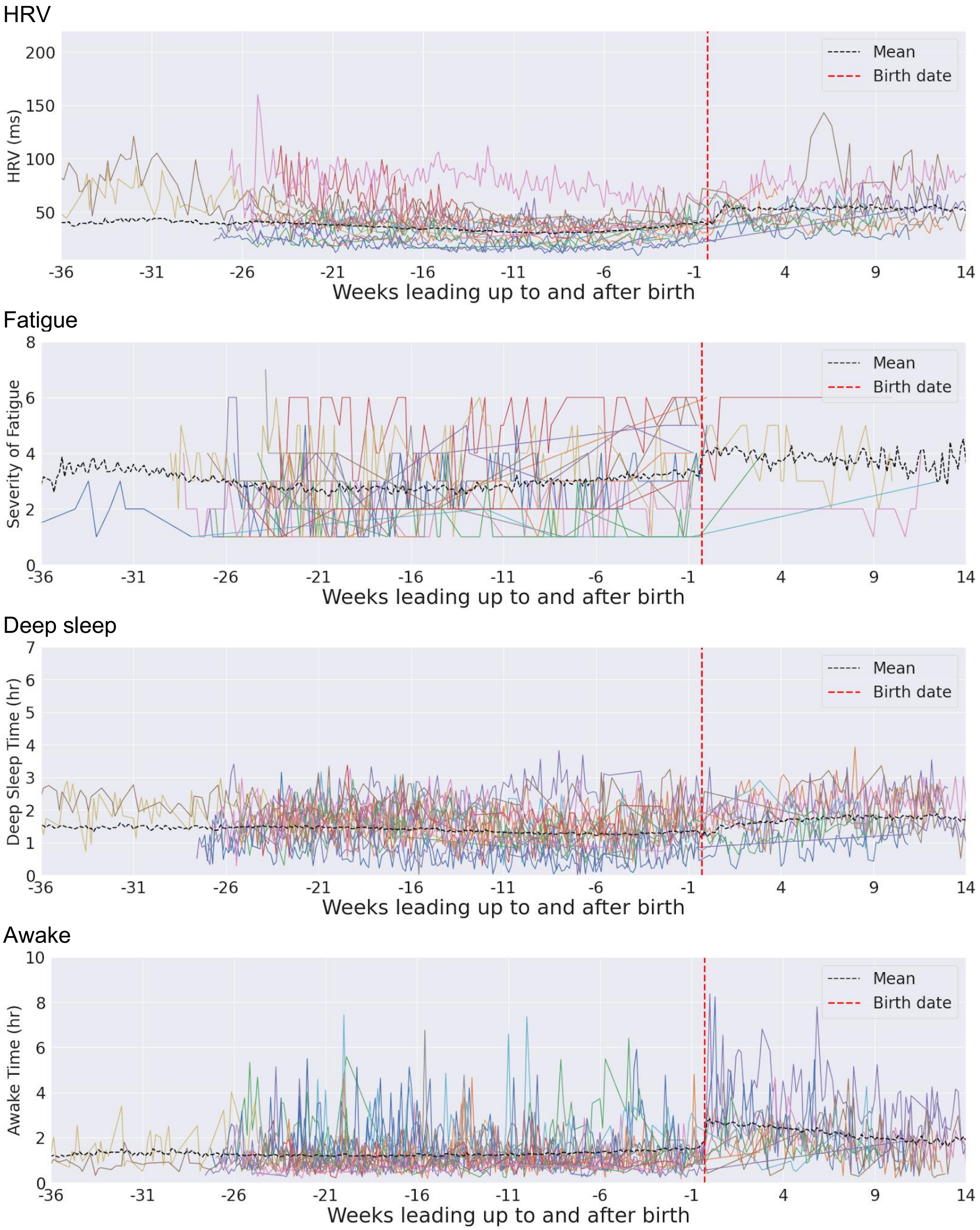


**Fig. S3. Spaghetti plots of four features for all healthy groups during pregnancy.**

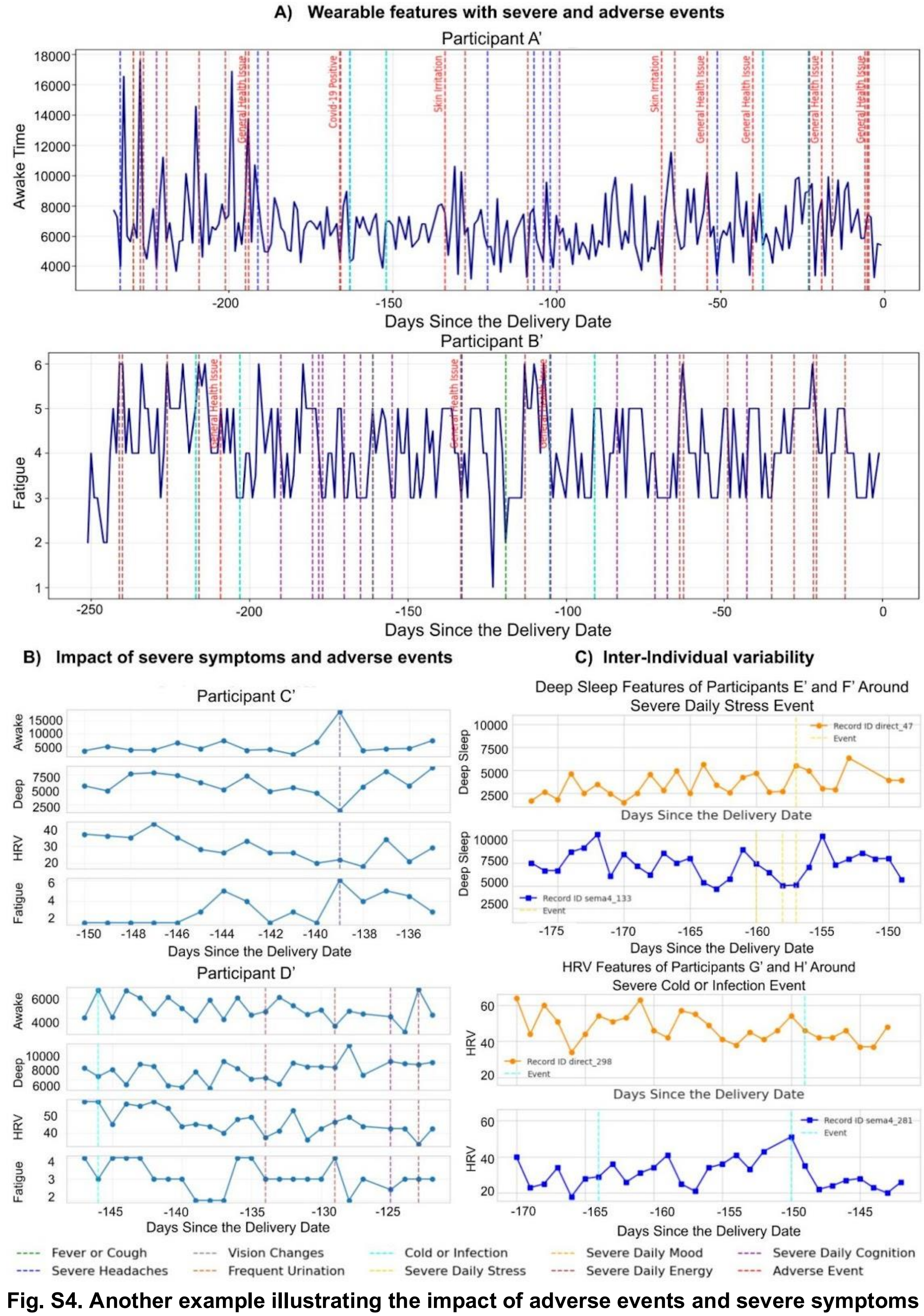


**Fig. S4. Another example illustrating the impact of adverse events and severe symptoms on fatigue, deep sleep, awake time, and HRV features.**

**Supporting Information Tables**

**Table S1. Daily activity survey**

| | Question | Answer options |
|---|---|---|
| Stress | What is your current stress level? | Very stressed, stressed, neutral, calm, very calm, no answer |
| Energy | Please assess your current energy level. | Very Energetic, Energetic, Neutral, Low Energy, Very Low Energy, no Answer |
| Cognition | What is your brain capable of performing right now? | All complex tasks, some complex tasks, moderate tasks, all simple tasks, some simple tasks, no answer |
| Mood | Please assess your mood. | Very positive, positive, neutral, negative, very negative, no answer |

**Table S2. Severe symptom and complication survey. The following survey asks about each participant's symptoms in the PAST 2 WEEKS. There are 7 yes/no questions, each taking 30 seconds to answer.**

| | Questions |
|---|---|
| Severe Cold Infection | Have you had any cold or infection of any kind in the past 2 weeks? |
| Severe Fever Cough | Have you had any fever, sore throat or cough in the past 2 weeks? |
| Frequent Burning Urination | Have you had burning or frequent urination in the past 2 weeks? |
| Severe Headache | Have you experienced severe headaches in the past 2 weeks? |
| Severe Vision Change | Have you experienced changes in vision in the past 2 weeks? |

**Table S3. Summary of Adverse Events. Final labeling used for grouping free-text adverse event notes and the count of these events across 275 individuals.**

| Health Issue | Count |
|---|---|
| General Health Issue | 132 |
| Covid-19 Positive | 35 |
| Pain | 34 |
| Skin Irritation | 11 |
| Fever & Body Aches | 10 |
| Nausea | 7 |
| Fatigue | 7 |
| Fall | 6 |
| Hypertension | 4 |
| Hyperglycemic | 4 |
| Emergency Visit | 4 |

**Table S4. Frequency of severe symptoms and adverse events reported. The table includes a comprehensive list of events, including Severe Daily surveys for very high stress, very low energy, low cognition in all tasks, and very negative mood, as well as Severe Fever/Cough, Severe Headache, Severe Cold/Infection, Severe Vision Change, and Adverse Events (AE).**

| | Severe Daily Stress | Severe Daily Energy | Severe Daily Cognition | Severe Daily Mood | Severe Fever Cough | Severe Headache | Severe Cold Infection | Frequent Burning Urination | Severe Vision Change | AE |
|---|---|---|---|---|---|---|---|---|---|---|
| count | 201 | 990 | 2603 | 143 | 165 | 158 | 170 | 57 | 39 | 148 |

**Table S5. Summary statistics (mean, variance, minimum, and maximum) of women's demographics. Parity refers to the number of previous births.**

| Pre Pregnancy demography attributes | mean | std | min | max |
|---|---|---|---|---|
| Age | 36.52 | 4.28 | 25 | 52 |
| Weight (lbs) | 159.68 | 35.05 | 104 | 286 |
| BMI | 26.46 | 5.81 | 17.93 | 46.15 |
| Parity | 0.32 | 0.60 | 0 | 2 |

Participation from non-white women is low, accounting for less than 20%. For example, only 6% of participants are Asian, and 5% are Black women.

**Table S6. Summary of Pre-pregnancy Conditions.**

| Pre-pregnancy Conditions | Mean | Std | Count |
|---|---|---|---|
| Allergies | 0.393162 | 0.490553 | 117 |
| Anxiety disorder | 0.364407 | 0.483316 | 118 |
| Autoimmune disorder | 0.161017 | 0.369114 | 118 |
| Blood Clotting Disorder | 0.008403 | 0.091670 | 119 |
| Cancer | 0.025210 | 0.157426 | 119 |
| Diabetes | 0.033898 | 0.181739 | 118 |
| Eating Disorder | 0.072165 | 0.260105 | 97 |
| Heart Disease | 0.008403 | 0.091670 | 119 |
| Hypertension | 0.059322 | 0.237234 | 118 |
| Kidney Disease | 0.016807 | 0.129090 | 119 |
| Mood disorder (e.g., Depression, Bipolar disorder) | 0.277311 | 0.449564 | 119 |
| Neurologic disorders (e.g., epilepsy, multiple sclerosis) | 0.016807 | 0.129090 | 119 |
| Psychotic Spectrum Disorder (e.g., Schizophrenia) | 0.016807 | 0.129090 | 119 |
| Pulmonary (e.g., Asthma, COPD) | 0.142857 | 0.351407 | 119 |
| Sleep disorder | 0.084746 | 0.279691 | 118 |
| Thyroid dysfunction | 0.151261 | 0.359818 | 119 |

**Table S7. Physiological Trends and Variability in Pregnancy: HRV, Fatigue, Sleep and Awake Features. The first column represents the percentage of individuals within the range of aggregate outcomes. A lower percentage in the first column and a higher CV indicate greater variability relative to the mean, reflecting less consistency in the outcome.**

| | Physiological Trends in Pregnancy (%) | Absolute Coefficient of Variation (CV) |
|---|---|---|
| HRV Inflection Point | HRV Inflection Point (Week 33): 4.76% | 14.24% |
| Fatigue Trough | Fatigue Trough (Week 21): 5.64% | 38.37% |
| Fatigue Peak | Fatigue Peak (Weeks 7 & 8): 9.69% | 67.42% |
| Sleep Slope | Sleep Slope (-0.02): 21.82% | 192.40% |
| Awake Slope | Awake Slope (0.02): 17.92% | 211.81% |
